# Improving epileptogenic zone estimation using Bayesian inference on neural field models

**DOI:** 10.1101/2023.10.01.23296377

**Authors:** Anirudh Nihalani Vattikonda, Marmaduke M. Woodman, Jean-Didier Lemarechal, Daniele Daini, Meysam Hashemi, Fabrice Bartolomei, Viktor Jirsa

## Abstract

Epilepsy remains a significant medical challenge, particularly in drug-resistant cases where surgical intervention may be the only viable treatment option. Identifying the epileptogenic zone, the brain region responsible for seizure initiation, is a critical step in surgical planning. Combining dynamical system models and the neuroimaging data of epileptic patients in a Bayesian framework has previously been shown to be a promising approach to identify the epileptogenic zone. However, previous studies employed coupled neural mass models to describe the whole brain seizure dynamics and hence could only provide a highly coarse spatially estimate of the epileptogenic zone. In this study we propose an extension of the Bayesian virtual epileptic patient framework to a neural field model which could improve the spatial resolution by several orders. Performing model inversion using neural field models is a challenging task since: (i) it is computationally expensive to compute gradients over a neural field simulation and (ii) parameter space can be very high dimensional. We demonstrate that using pseudo-spectral methods and spherical harmonic transforms it is feasible to perform Bayesian model inversion on a neural field extension of the reduced Epileptor model. We found that the neural field extension not only improves the spatial resolution but also significantly reduces the number of false positives.

## Introduction

Treatment for focal drug resistant epilepsy typically involves surgical resection of brain regions, called Epileptogenic Zone (EZ), which are hypothesized to be inducing seizures. However, in only about 60% of the cases are the patients seizure free post surgery [1]. Recently, several studies have proposed various methods to improve the presurgical estimation of EZ which could thereby improve surgical outcomes.

Epileptogenicity Index (EI) [2] uses spectral and temporal information to define a quantitative measure of the degree of epileptogenicity of brain regions recorded through stereotactic EEG (SEEG). Epileptogenicity Maps (EM) [3] provide anatomical map of seizure onset and propagation using spectral analysis of fast activity ranging from 60 to 100 Hz. Connectivity Epileptogenicity Index (cEI) [4] combines a directed connectivity based measure with EI to improve EZ estimation in seizures with slow onset patterns. Epileptogenicity Rank (ER) [5] is a modified EI measure which utilized temporal, spatial and energy ratio during ictal discharges to estimate EZ. These methods are purely data driven i.e. the EZ is estimated based on quantitative and/or statistical analysis of SEEG recordings of epileptic patients.

Alternatively, combining dynamical system models of epileptic seizures with Bayesian inference techniques has been demonstrated to be a promising approach to improve the presurgical estimation of the EZ. Such approaches mainly involve (i) defining a probabilistic model of seizure dynamics and (ii) inferring parameters of the dynamical model using a Bayesian framework. Dynamical causal modeling (DCM) [6] is a popular framework in Neuroscience that demonstrated the usefulness of such an approach in understanding various cognitive processes such as vision [7, 8], language [9, 10], decision making [11, 12] etc. In the context of epilepsy, DCM has been used to study: seizure propagation pathways from simultaneous EEG-fMRI data [13], modulations in synaptic efficacy between regions during seizure onset[14], concurrent changes in synaptic coupling at macro and meso-scales from calcium imaging data in zebrafish [15].

DCM is primarily used to study the mechanisms of effective connectivity using biophysically plausible neuronal models. Virtual epileptic patient (VEP) [16, 17] proposed a similar framework to DCM but principally aimed towards building personalized epilepsy models using a phenomenological model of epileptic seizure dynamics. Following studies have demonstrated the efficacy of VEP approach in identifying EZ using point estimates [18] and fully Bayesian posterior estimates [19] on a retrospective patient cohort. In VEP, the seizure dynamics are described using a neural mass model (NMM) called Epileptor [20, 21]. NMMs can describe the dynamics of the whole brain by considering brain as a network of discrete interacting neuronal populations, where the interactions are typically constrained by the structural connectome (SC) [22]. NMMs have been extensively used to understand the macroscopic brain dynamics in healthy [23, 24, 25, 26, 27] and disease conditions [28, 29, 30, 21]. One of the key limitations of the previous VEP studies is the highly coarse spatial resolution owing to the use of neural mass model [31]. In order to address this, here we propose a neural field extension of VEP. While NMMs describe dynamics across time, neural field models (NFM) describe the dynamics across space and time, allowing for infinite spatial resolution in theory. Moreover, compared to NMMs, NFMs were shown to better explain empirically observed LFP data [32]. However, several challenges exist in performing Bayesian model inversion of NFMs: (i) they are computationally very expensive to simulate at the whole brain level, (ii) model inversion requires computing gradients which is computationally much more intensive than the simulation and (iii) dimensionality of the parameter space scales linearly with the spatial resolution.

Mapping the cortical surfaces onto a spherical surface and using a pseudo spectral approach for numerical integration was recently shown to reduce the computational cost of neural field simulations by several orders of magnitude [33, 34]. In this study, using the pseudo spectral approach and reparameterizing the parameter space into spherical harmonic mode coefficients, we first demonstrate the plausibility of Bayesian model inversion of a neural field extension of the reduced Epileptor model. To our knowledge this is the first study to demonstrate model inversion of a NFM at whole brain level. Using synthetic data, robustness of the inversion across different resolutions and hyper-parameters is systematically tested. Finally, we study the accuracy of the field extension of VEP in estimating EZ on a retrospective patient cohort and compare it with the previous NMM variant of VEP.

## Methods

### Patient data

To study the accuracy of the model, it is tested against a retrospective patient cohort of 12 patients. The dataset used in this study is the same as the one used in a previous VEP study [18, 31, 19]. The dataset includes invasive and non-invasive multimodal neuroimaging data such as T1-MRI, diffusion MRI and SEEG. For more details of the data acquisition process please refer to [31]. Informed written consent was obtained for all patients in compliance with the ethical requirements of the Declaration of Helsinki, and the study protocol was approved by the ethics committee Comité de Protection des Personnes sud Méditerranée 1. For each patient with a known surgical outcome, a connectome is generated by running the reconstruction pipeline at high spatial resolutions with brain surface modeled as a triangular mesh containing 327684 vertices. Some of the patients are excluded due to issues with running the reconstruction pipeline at high spatial resolutions. Out of the 25 patients in the original dataset, the reconstruction pipeline ran successfully for 12 patients at high spatial resolution. These 12 patients are used for empirical validation in this study. Among the 12 patients, 7 are seizure free post surgery with their surgical outcome rated as Engel score I. In the remaining 5 patients surgery failed to achieve seizure freedom and their outcomes are classified as either Engel score II, III or IV. More details of the patients such as age, gender, Engel scores and inclusion/exclusion criteria are provided in Table 1. Patient IDs in Table 1 are not known to anyone outside the research group and does not reveal the identity of the patient to anyone outside the research group.

**Table 1:**
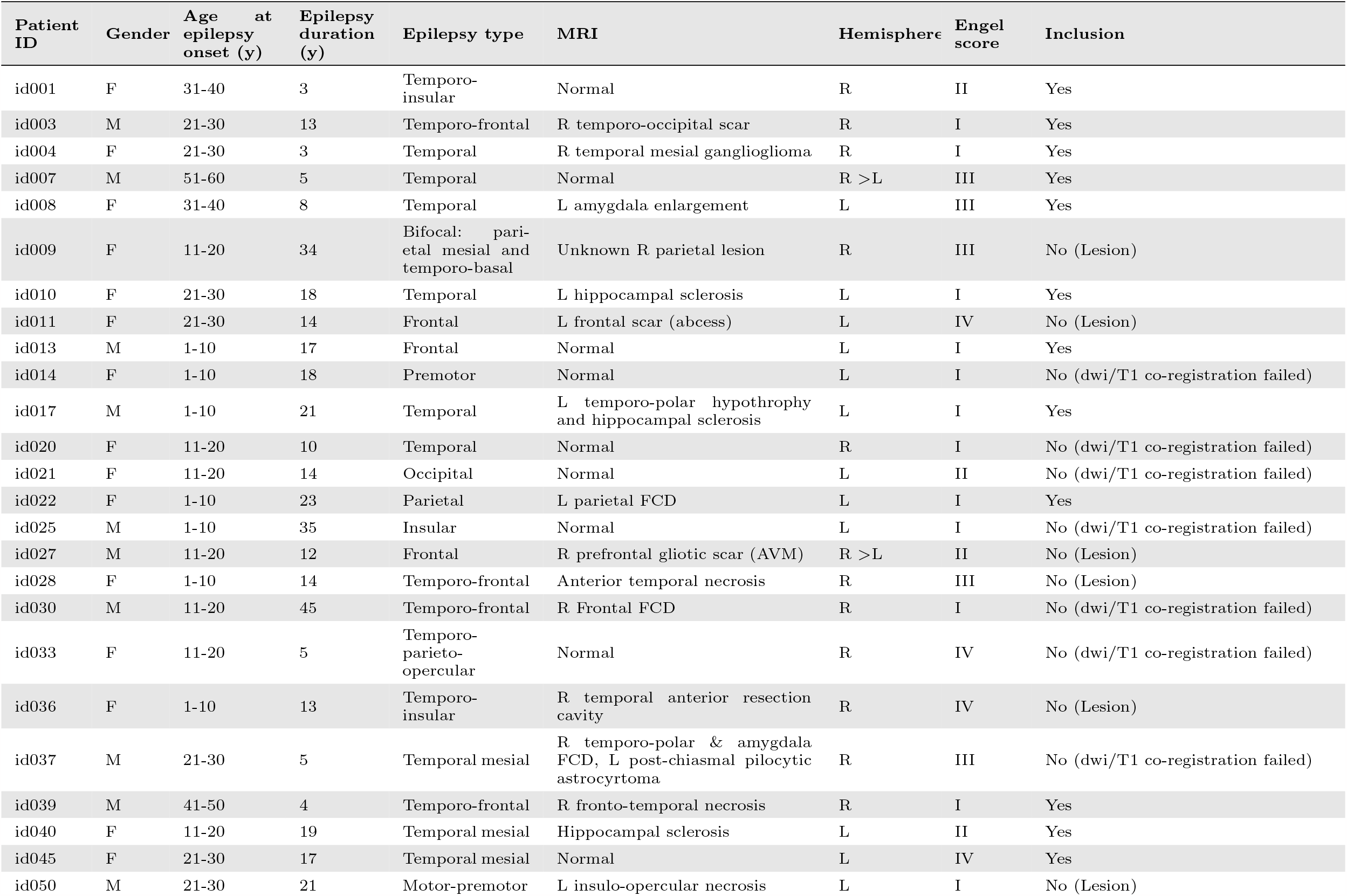
Details of the retrospective patient dataset. Abbreviations: AVM=arteriovenous malformation; FCD=focal cortical dysplasia; L=left; R=right.

#### Connectome

For each patient, using the reconstruction pipeline, a connectome is generated by parcellating the brain according to VEP atlas [35] consisting of 162 regions with 72 cortical and 9 subcortical regions per hemisphere. Similar to other whole brain studies using neural mass models [23, 25, 22], the VEP framework [21, 16, 17] uses the connectome to define the long range influence between brain regions. In NMM models of whole brain activity, the influence of one region on other is constrained by scaling the activity of a region by the connectivity. However, in this study, since a neural field model is used, long range coupling between two distinct regions is defined using the average activity of all vertices in a region with the connectome constraining the influence of that region on others.

#### Forward model

Neural activity at vertices is projected to the activity at the sensors using a so called Gain matrix (*G*) [31, 18]. Assuming that the activity decays with square of distance from the source, the gain coefficient mapping activity at source vertex *i* to sensor *j* is estimated as:

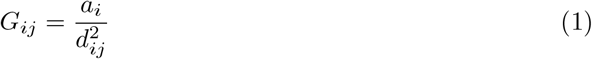

where, *a*_*i*_ is the area of vertex *i* obtained by summing up one-third of the area all neighboring triangles. In this study, we use a pseudo-spectral method to compute numerical solutions of the neural field model. The pseudo-spectral method entails mapping the inflated spherical pial surfaces obtained from FreeSurfer [36, 37] to a regular grid on the spherical surface. The gain matrix corresponding to the regular spherical grid is estimated as the sum of nearest vertices on the irregular grid i.e.

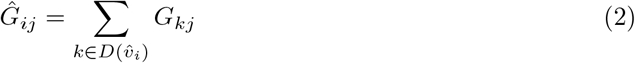

where 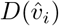 represents all the vertices on the irregular spherical grid whose nearest neighbor is the vertex 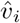 on the regular spherical grid.

#### Resection Masks

Post surgical MRI is manually inspected to identify the brain regions in the VEP parcellation that are resected by the surgery. A resection mask is created as a binary vector with as many elements as the number of regions in the VEP parcellation. The resection mask (*M*) is defined as:

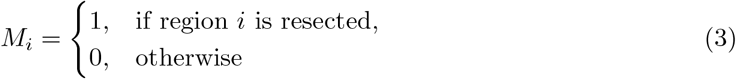

where *i* ∈ (1, 162) indexes one of the brain regions of the parcellation.

Since a neural field model is used in this study, the model predicted EZ are described at a high spatial resolution with 32776 vertices per hemisphere. For comparison with the empirical outcomes, the resection masks at parcellation level (Eq. 3) are mapped to high resolution (*M*^*H*^) as follows:

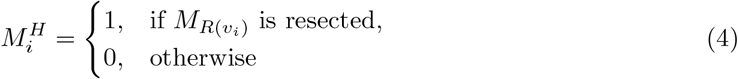

where, *i ∈* (1, 65554), *v*_*i*_ represents vertex *i* and *R*(*v*_*i*_) represents the index of brain region in the VEP parcellation that vertex *v*_*i*_ belongs to.

### Synthetic data

To study the accuracy of the EZ predictions by comparing them with ground truth two synthetic datasets are generated using the connectome and gain matrices of the patients *id*004 and *id*022. EZ are defined by choosing regions close to the implanted electrodes. Specifically, for the patient *id*004 EZ consisted of three regions: Right-Temporal-pole, Right-T2-posterior and Right-Thalamus. EZ for *id*022 consisted of four regions: Left-Superior-parietal-lobule-P1, Left-Intraparietal-sulcus, Left-Postcentral-gyrus, Left-Postcentral-sulcus. SEEG electrode implantation, centers of the brain regions in EZ, structural connectome and gain matrix of patient *id*004 and *id*022 are shown in supplementary Fig. 1 and supplementary Fig. 3 respectively. For all the vertices belonging to regions in EZ the tissue excitability parameter (*x*_0_) is set to *−*1.8 and for the rest of the vertices *x*_0_ = *−*3.0. Source activity for this spatial map of excitability is obtained by numerically solving the neural field extension of the 2D Epileptor model using a pseudo spectral method. Initial values of the fast (*x*) and slow (*z*) variables are set to *−*2.0 and 5.0 respectively.

### Feature extraction

The raw SEEG data is preprocessed to extract SEEG log power. Preprocessing involves high pass filtering raw SEEG data from 10 Hz, computing the power over a sliding window, applying a log transformation and finally a low pass filter of 0.05 Hz is applied for smoothing out any short spikes in the data. In order to reduce the computational cost of fitting, the SEEG log power time series is down sampled to 300 time points. Data augmentation is a technique in machine learning to improve optimization when the observations are sparse. A common technique for data augmentation is to increase the number of observations by adding noise to the observed data. In this study, each time series is augmented by generating 50 more time series by adding Gaussian noise with mean zero and standard deviation of 0.1.

### Dynamical Model

Epileptor is a phenomenological model developed to describe the predominant dynamical characteristics of focal epileptic seizures [20]. Later, studies have extended the Epileptor model to study the mechanisms of seizure recruitment and propagation through neural mass models [21] and neural field models [38]. The Epileptor model consisted of five state variables with two variables capturing fast discharges, two variables for spike and wave events and one variable accounting for extracellular effects and governing autonomous evolution between interictal and ictal phases. Taking advantage of time scale separation the Epileptor model could be reduced to two variables with the one slow variable similar to the full model and one fast variable which could be interpreted as proxy to transients in power dynamics between interictal and ictal phases. Previously, by embedding the 2D Epileptor neural mass model into a Bayesian framework [17, 18, 31, 19, 16] it was demonstrated that model inversion could be performed to generate individualized models of seizure recruitment and propagation which can improve presurgical identification of EZ. In this study, we propose a neural field extension of the reduced Epileptor model aimed towards improving the spatial resolution of the EZ predicted by the BVEP framework [17, 18]. The dynamics of the two dimensional Epileptor neural field model at position *y* and time *t* is defined as follows:

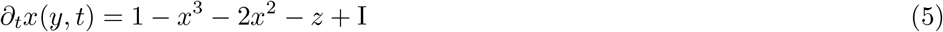

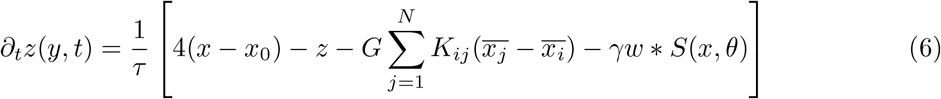

where *G* represents the global coupling i.e. a scaling factor for long range connectivity *K*. The indices *i* and *j* represent the index of the brain regions in VEP parcellation, with *i* indexing the region at position *y* and *j* is a dummy index variable looping across all brain regions. 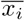 represents the average value of state variable *x* in region *i*. The term *w S*(*x, θ*) represents the local or homogeneous coupling with the operator representing a spatial convolution. It is defined as:

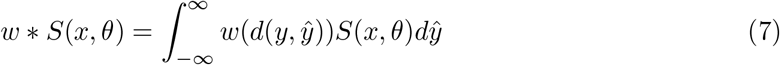

where *d*(., .) is the distance along the surface between two points, *S* is a sigmoid function i.e. *S*(*q, θ*) = 1*/*(1 + *e*^*−*(*q−θ*)^) and *w* is a short ranged function representing local connectivity kernel typically defined as a Laplacian i.e. *w*(*y*) = *e*^*−*|*y*|^*/*2. In this study unless otherwise specified *γ* = 1 and *θ* = *−*1. In computing numerical solutions of neural field models the homogeneous connectivity was demonstrated to be a computational bottleneck [33]. Performing model inversion entails computing gradients over the generated solutions. Typically these gradients are computed using automatic differentiation [39] which builds a computational graph of all the operations or transformations involved in mapping a particular parameter configuration of the neural field model (Eq. 6) to the SEEG data features. Computationally intensive functions such as the homogeneous connectivity term in Eq. 7 can lead to prohibitively high memory usage in computing gradients. To avoid this, in this study, we use a pseudo-spectral method to estimate the homogeneous connectivity.

### Pseudo-spectral method

For the family of neural field equations that are generally described as:

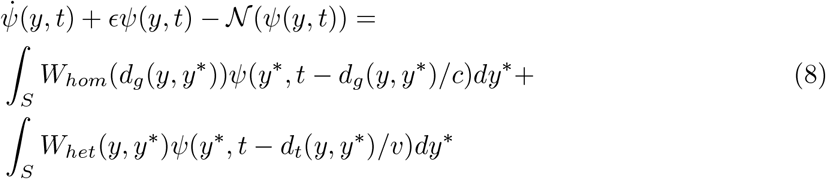

where, *ψ* represents the state vector, *𝒩* is a function representing the non-linear contributions, *d*_*g*_ and *d*_*t*_ represent distance along the surface between two points and length of the fiber tracts connecting two points respectively. *W*_*hom*_ and *W*_*het*_ are the homogeneous or short range and heterogeneous or long range connectivity kernels. The parameters *c* and *v* denote transmission speeds. It was shown that by mapping the high resolution inflated surfaces obtained from FreeSurfer [36, 37] onto a unit sphere, neural field solutions can be obtained through a pseudo spectral method [33] using mode decomposition on the spherical surface. Specifically, it was demonstrated that when the homogeneous connectivity kernel is a Laplacian the homogeneous connectivity could be approximated as:

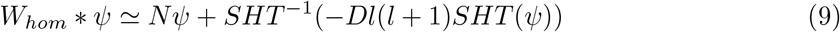

where *N* is a normalization coefficient, *D* is a diffusion coefficient and *SHT* indicates spherical harmonics transform. In approximating the homogeneous connectivity as given in Eq. 9, using the fact that the homogeneous kernel (Laplacian here) is a short ranged function, Daini and Jirsa [33] found *N* and *D* to be:

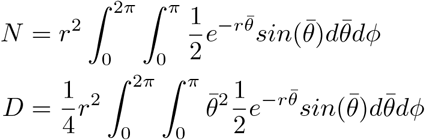

which for *r* = 100 mm are *N* ≃ 3.14128 and *D* ≃ 0.00047108. Computing the homogeneous connectivity using Eq. 9 requires performing spherical harmonic transform, in this study, unless otherwise specified the spherical harmonic mode coefficients are truncated up to a degree of 32.

#### Numerical Solution

To compute the trajectories of the neural field state variables governed by Eq. 6, we performed numerical integration using Euler’s method with a time step of 0.05 seconds for a total of 600 steps. Since the SEEG data features are subsampled to contain 300 time points, the time series of the state variables obtained from numerical integration is subsampled at every 0.1 second.

### Model inversion

In the Bayesian VEP framework [17, 18, 31] model inversion can be performed using either Maximum a posteriori (MAP), Variational inference or MCMC based sampling techniques over a posterior distribution:

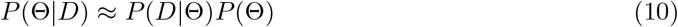

where, Θ is a vector containing parameters of interest and *D* is the observed data features such as the SEEG log power. The parameter vector *θ* contains, initial values of the state variables (*x*(*y, t*_0_), *z*(*y, t*_0_)), spatial map of tissue excitability (*x*_0_(*y*)), global coupling (*G*), time scale parameter *τ*, observation noise (*ϵ*) and two scalar auxiliary parameters amplitude (*α*) and offset (*β*) defining a global linear transformation of the model predicted SEEG log power. In the spatial domain, with a spatial discretization of 32768 vertices per cortical hemisphere, the dimensionality of the parameter space would be 196667. In order to reduce the dimensionality of the parameter space, all the spatial maps are parameterized using SHT up to a degree of *L*_*max*_. In this study we used *L*_*max*_ = 16, except while performing parameter sweep across *L*_*max*_ for sensitivity analysis, reducing the dimensionality of parameter space to 3527. The prior distributions over the above parameters are defined as:

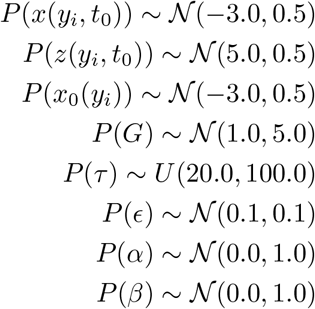

Taking advantage of prior knowledge of the parameter regimes of the dynamical model Eq. 6 for seizure recruitment and propagation all the priors with Normal distributions are bounded above and below. The upper and lower bounds used in this study are provided in table 2.

**Table 2:**
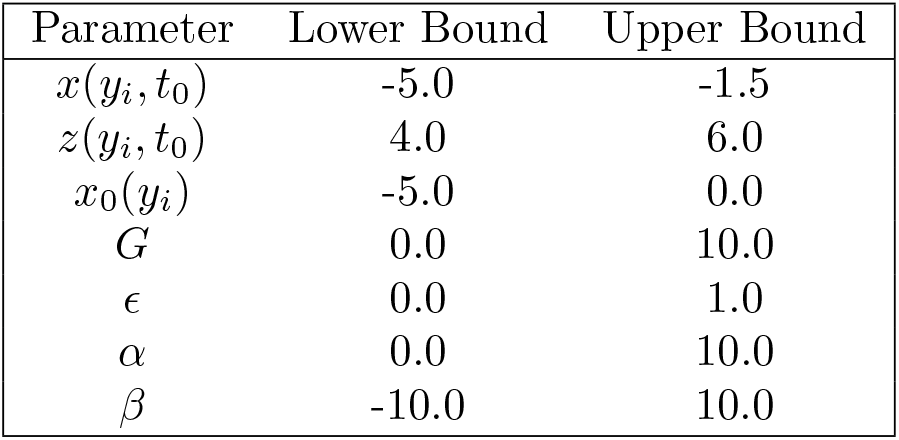
Upper and lower bounds for the inferred parameters.

| Parameter | Lower Bound | Upper Bound |
| --- | --- | --- |
| $x(y_i, t_0)$ | -5.0 | -1.5 |
| $z(y_i, t_0)$ | 4.0 | 6.0 |
| $x_0(y_i)$ | -5.0 | 0.0 |
| $G$ | 0.0 | 10.0 |
| $\epsilon$ | 0.0 | 1.0 |
| $\alpha$ | 0.0 | 10.0 |
| $\beta$ | -10.0 | 10.0 |

In this study, model inversion is performed using MAP. Adam optimizer [40] is used to perform optimization with a learning rate of 0.001 and 5000 steps as the termination criteria. Sensitivity analysis across different parameter sweeps are performed on CSCS high performance cluster where each node has the following configuraiton: Intel® Xeon® E5-2690 v3 @ 2.60GHz (12 cores, 64GB RAM) and NVIDIA® Tesla® P100 with 16GB graphics memory. Rest of the analysis is performed on a workstation with Intel® Core™ i9-10885H CPU @ 2.40GHz (16 cores, 64 GB RAM) and NVIDIA Quadro RTX 4000 GPU with 8GB graphics memory.

#### EZ Estimation

To identify the EZ from the inferred parameters, we used the seizure onset times similar to [18]. Onset times are estimated by finding the time instant where depolarization shift occurred in the model predicted activity of the fast variable (*x*). Next, all regions with onset times within a tolerance, *t*_*ϵ*_, of the earliest onset time, *t*_*λ*_, are classified as EZ. All other regions with onset times beyond *t*_*λ*_ + *t*_*ϵ*_ are classified as PZ.

## Results

### Validation on synthetic data

In order to compare the inferred EZ to the ground truth, first the inference accuracy is tested against synthetic data. Fig. 1a shows the tissue excitability parameter (*x*_0_) in ground truth and prediction while fitting against synthetic data described in methods. All three regions (RightTemporal-pole, Right-T2-posterior and Right-Thalamus) defined as part of EZ in the ground truth are accurately inferred to have higher excitability. In addition to these three regions, one other region (Right-T2-anterior) is inferred to have high excitability, even though, the fit to observations is good as shown in Fig. 1b. However, as shown in Fig. 2 this region is not recruited by the seizure. This demonstrates the structural degeneracy of the Epileptor model i.e. multiple configurations of the Epileptor model parameters can lead to similar source state dynamics. But this wouldn’t be a problem for the purpose of identifying EZ as it could be uniquely determined given the trajectory of source states.

**Figure 1:**
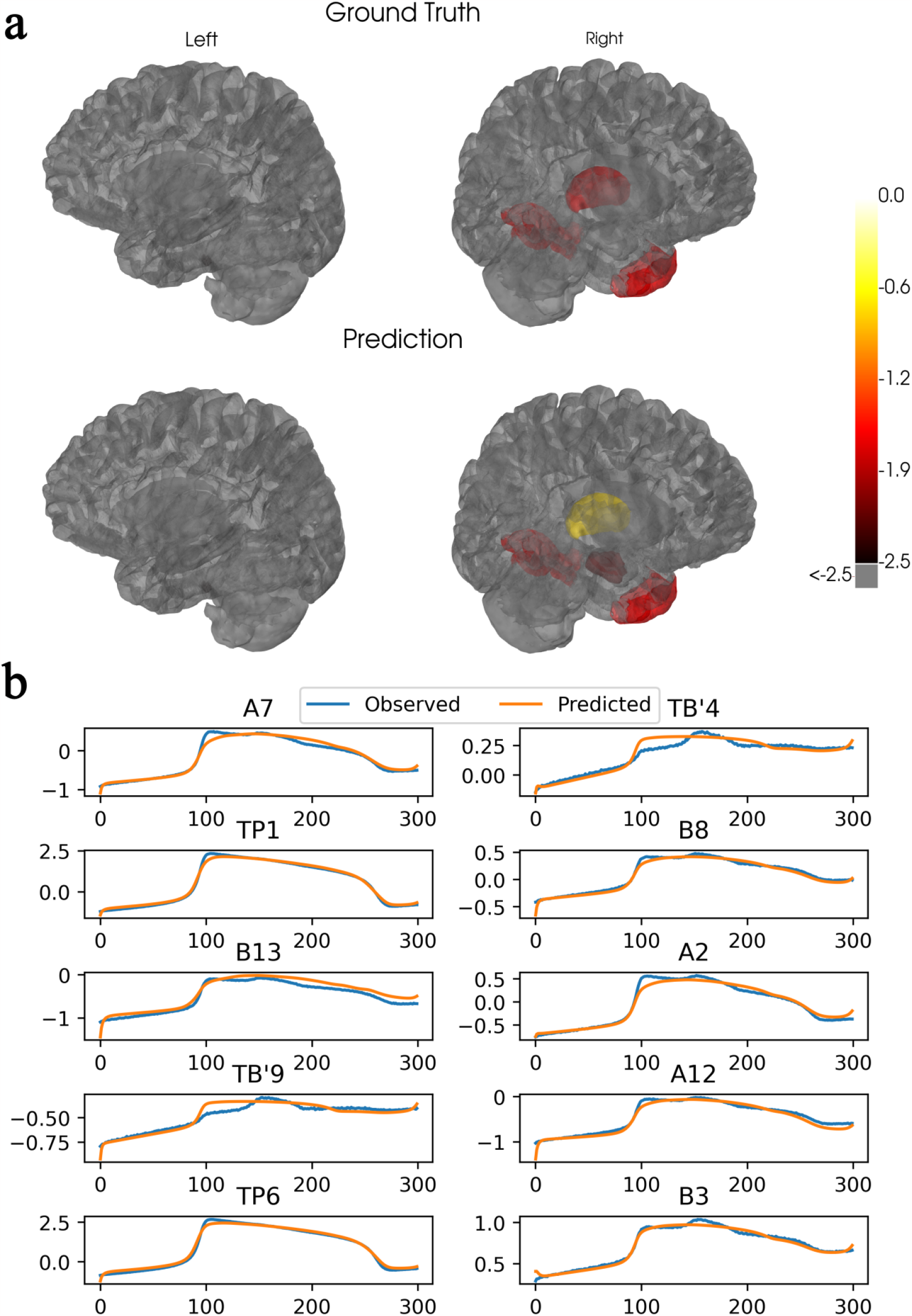
Inference accuracy on a synthetic dataset. (a) Comparison of the tissue excitability parameter (*x*_0_) between the ground truth i.e. synthetic data and the inferred values. (b) Goodness of fit between the observed and the predicted SEEG log power at 10 randomly chosen SEEG sensors

**Figure 2:**
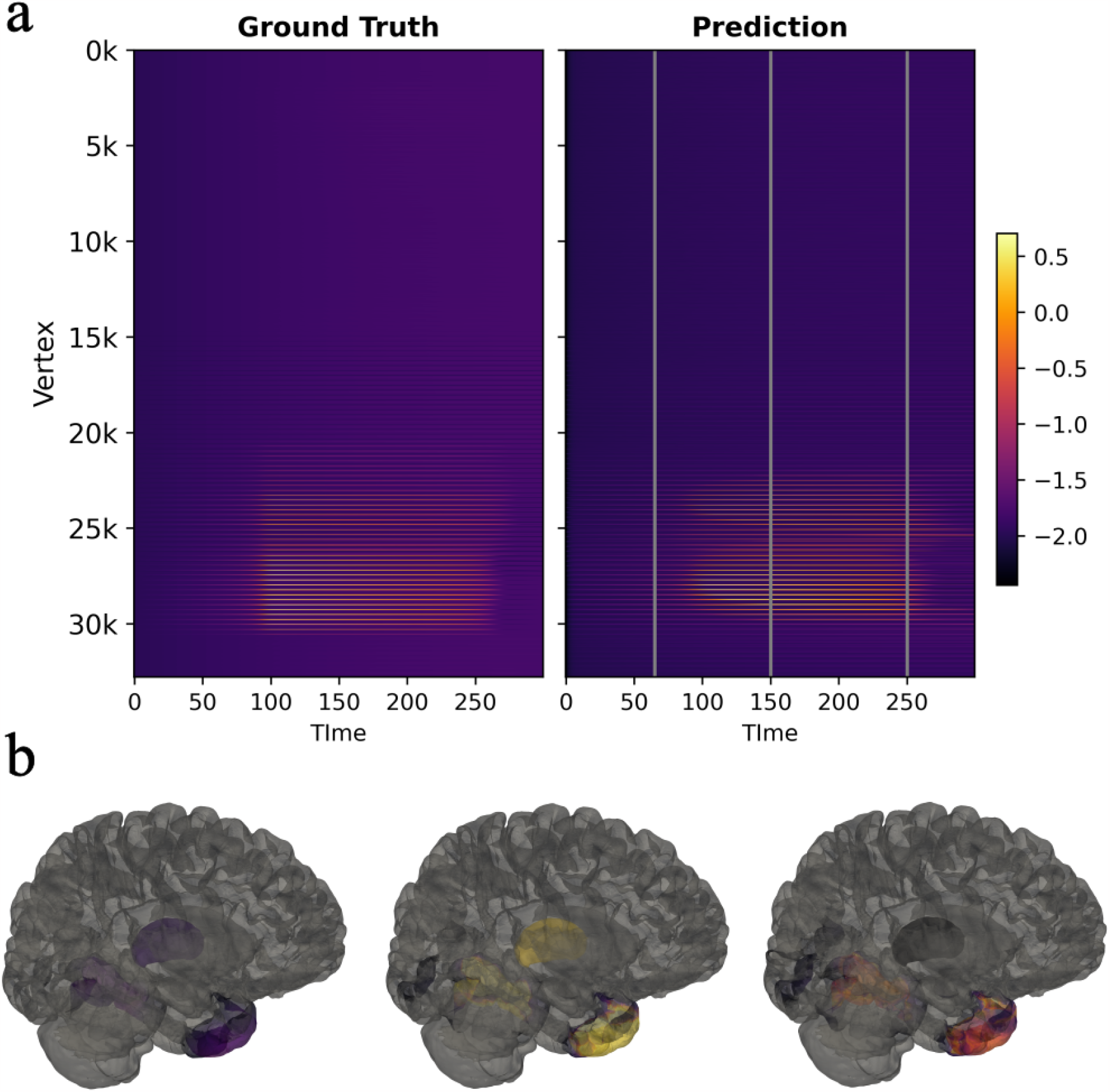
Comparison of source activity between ground truth and predictions. (a) Latent source activity obtained from the pseudo-spectral simulation of the neural field extension of 2D Epileptor model (left) and source activity obtained from a simulation of the same model using the model parameters estimated by performing model inversion using MAP (right). (b) inferred source activity at three different time steps, shown by gray vertical lines in “a”, illustrated on the brain surface. Color map is the same as in the one shown in “a”.

Next, we tested the robustness of the method to changes in spatial resolution, mode truncation and observation noise. We found that the method is robust across different spatial resolutions but sensitive to mode truncation and, as expected, to observation noise. As shown in Fig. 3a, both precision and recall of the inferred EZ remained above 80% across different spatial resolutions, ranging from 8192 vertices to 32,768 vertices. Accuracy of the inferred EZ is found to decrease when higher spherical harmonic modes i.e. *L*_*max*_ *>* 25 are used to represent the spatial maps of parameters (Fig. 3b). This could be because higher number of modes have higher expressive power i.e produce many distinct spatial maps which in turn would lead more local optimum. Interestingly, this would imply lower truncations not only help reduce the dimensionality of parameter space but also reduce the structural degeneracy in Epileptor model by constraining the spatial maps of parameters to those that have smooth spatial transitions. However, such smooth spatial maps would have the trade off that size of the inferred EZ could be underestimated to avoid false positives. To ensure that effect of hyperparameters and observation noise on accuracy are not specific to the EZ hypothesis, the same parameter sweeps are performed using bad EZ hypothesis. Their effects are found to be similar to the case with good EZ hypothesis (Fig. 3 d,e,f). Similar analysis performed on a different dataset is shown in supplementary Fig. 5.

**Figure 3:**
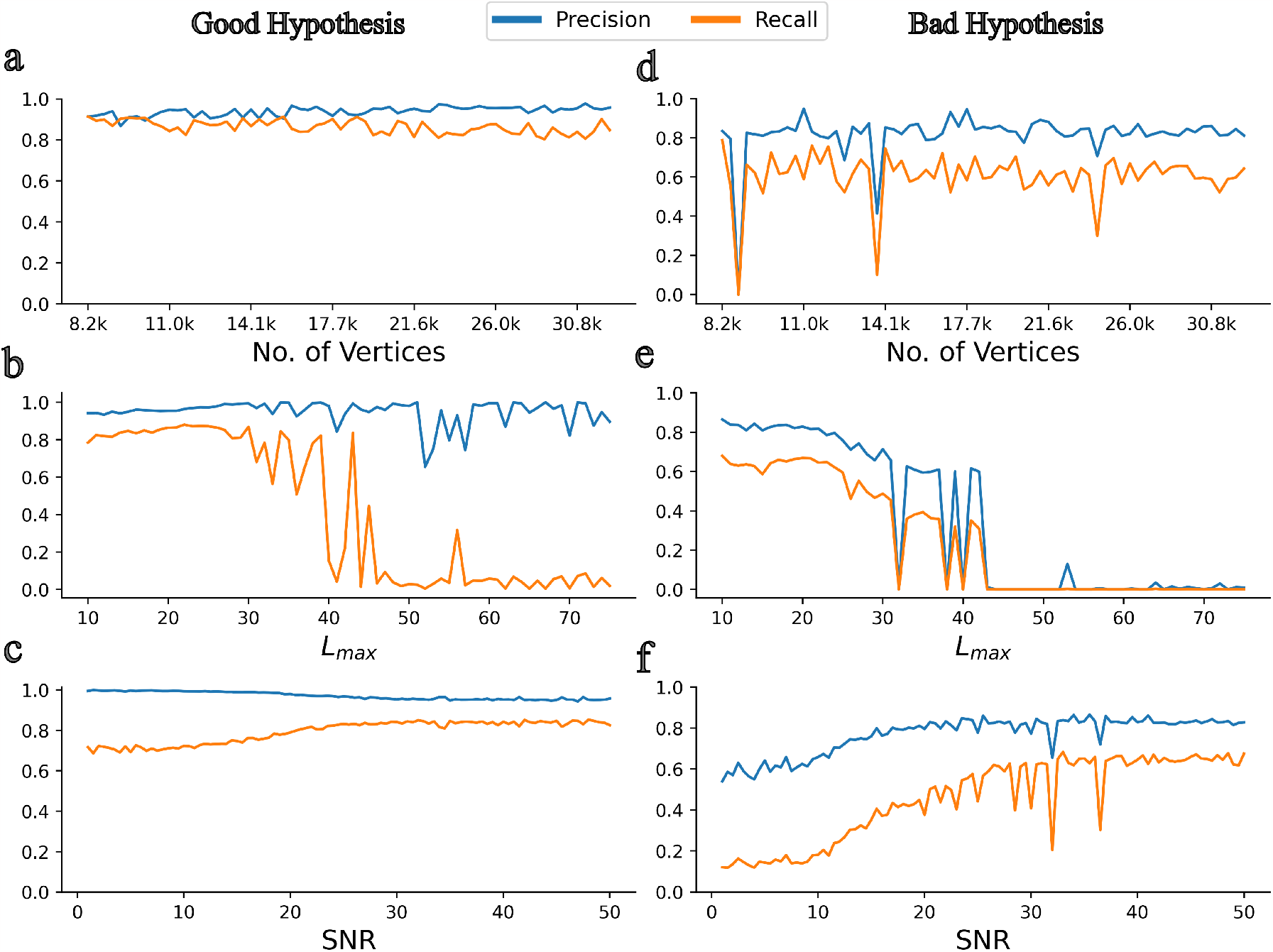
Sensitivity analysis across different hyper-parameters of VEP with neural field model (VEP-NFM) against synthetic data. (a) Precision and recall of VEP-NFM model across different spatial resolutions with a good EZ hypothesis (b,c) Same as “a” except across different mode truncation parameters and signal to noise ratio in the observed SEEG. (d,e,f) Same as “a,b,c” except with a bad EZ hypothesis.

### Validation on Empirical Data

Accuracy of the predicted EZ in empirical data is validated using a retrospective patient cohort consisting of 12 patients (see table). Out of the 12 patients, 7 patients are seizure free post surgery and the remaining 5 are not. Precision and recall of the predicted EZ compared to resection masks created from post surgical MRI is shown in Fig. 4. Precision/recall across seizures in each patient is shown in Fig. 4a. In patients who are seizure free post surgery i.e. Engel score I, on average, precision and recall are 0.85 and 0.25 respectively. In patients who are not seizure free post surgery, average precision recall are 0.58 and 0.17 respectively. As evident from the precision, in patients where surgery was successful in achieving seizure freedom, the number of false positives are much less compared to patients who are not seizure free post surgery. This would imply that the predicted EZ matches with the resected regions in patients where the surgery was successful but showed a mismatch in patients where surgery failed.

**Figure 4:**
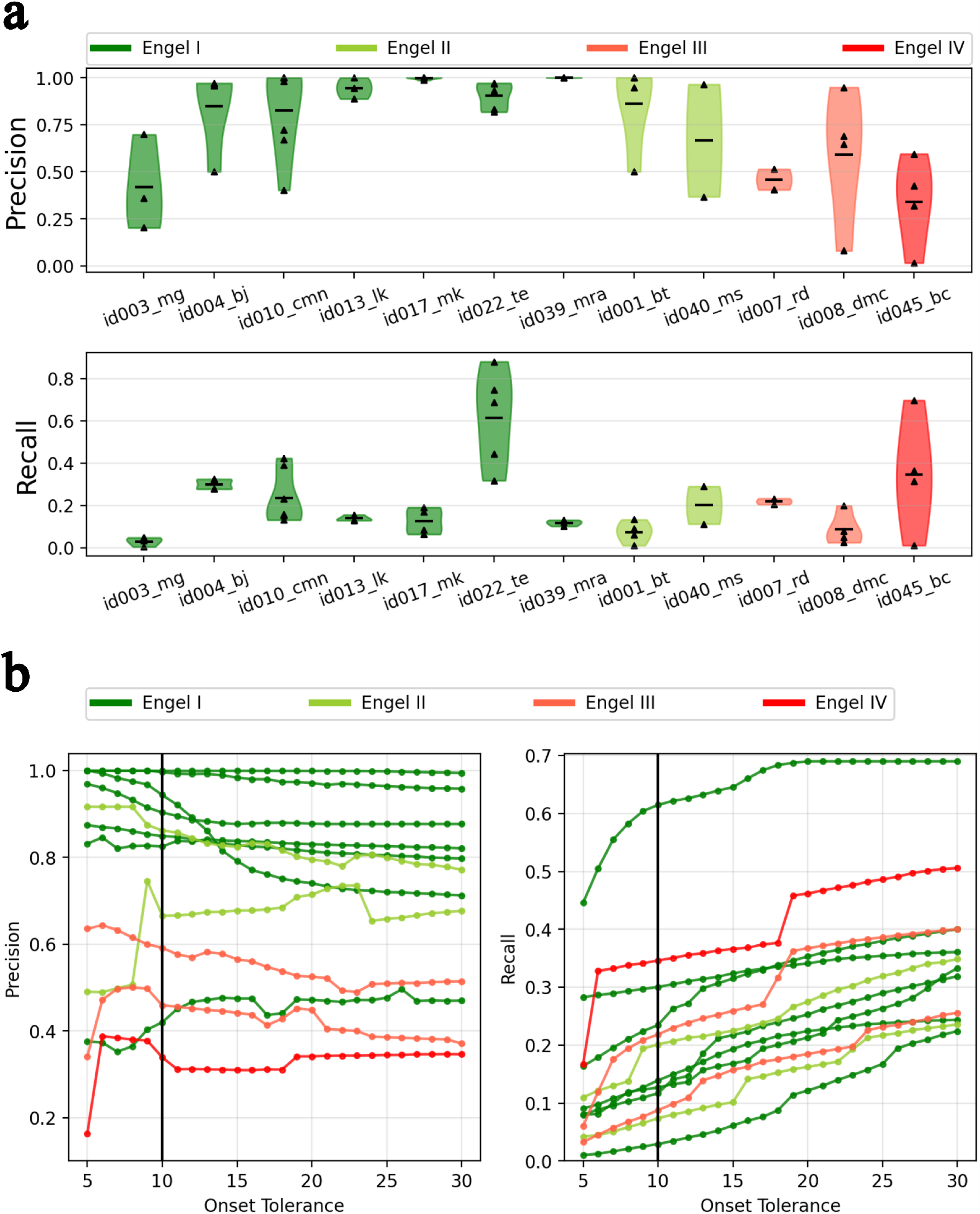
Analysis of accuracy of predicted EZ in a retrospective patient cohort.(a) Distributions of precision and recall across seizures in each patient. Precision and recall are computed by comparing the predicted EZ against EZ defined by post surgical MRI and setting the onset tolerance threshold to 10 seconds. (b) Average precision and recall, averaged across seizures, in each patient across different onset tolerance thresholds.

Note that the low recall values does not necessarily imply that most of the resected regions are predicted to be healthy. We hypothesize this could be because (i) some of the vertices classified as EZ in resection could be predicted as PZ based on onset tolerance threshold and (ii) the resection masks are created by mapping low resolution (163 nodes) parcellation masks to high resolution (32768 vertices), but the predicted EZ, even if it includes all the distinct resected regions, it may only include a fraction of the vertices from each region. To check the influence of onset tolerance threshold, precision and recall are computed at different thresholds ranging from 5 seconds to 30 seconds. Average precision and recall, across seizures, of predicted EZ in each patient at different thresholds is shown in Fig. 4b. Increasing the onset tolerance from 10 seconds to 30 seconds, resulted in almost two fold increase in recall. Average recall, per group, increased from 0.25 to 0.39 and 0.17 to 0.34 in seizure free and non seizure free patients respectively. While, average precision decreased slightly from 0.85 to 0.81 and 0.58 to 0.53 in seizure free and non seizure free groups respectively. The effect of constructing high resolution resection masks, from information at the parcellation level, on recall can be investigated by computing the recall at parcellation level. Predicted EZ at high resolution are mapped to parcellation level by thresholding on the percentage of vertices of a region in the predicted EZ. Accuracy of predicted EZ in each patient when mapped to parcellation level is shown in Fig. 5. Considering regions with at least 20% of its vertices to be part of EZ and at an onset tolerance of 30 seconds, average recall increased to 0.50 and 0.41 in seizure free and non-seizure free groups respectively. At the same thresholds, precision at the parcellation level is found to be almost same as that observed at high resolution, 0.81 and 0.52 specifically. The results observed in 4b and 5 provide evidence to our hypothesis that the low recall observed in 4 is a consequence of either thresholding or mapping information at parcellation level to high resolutions. These effects are then summarized by performing a parameter sweep over both the thresholds of onset tolerance and percentage of vertices. Average precision and recall at the parcellation level across different thresholds is shown in Fig. 6. As hypothesized, the recall increases as either onset tolerance is increased or by mapping EZ to parcellation level with lower fractions of vertices as inclusion criteria for EZ.

**Figure 5:**
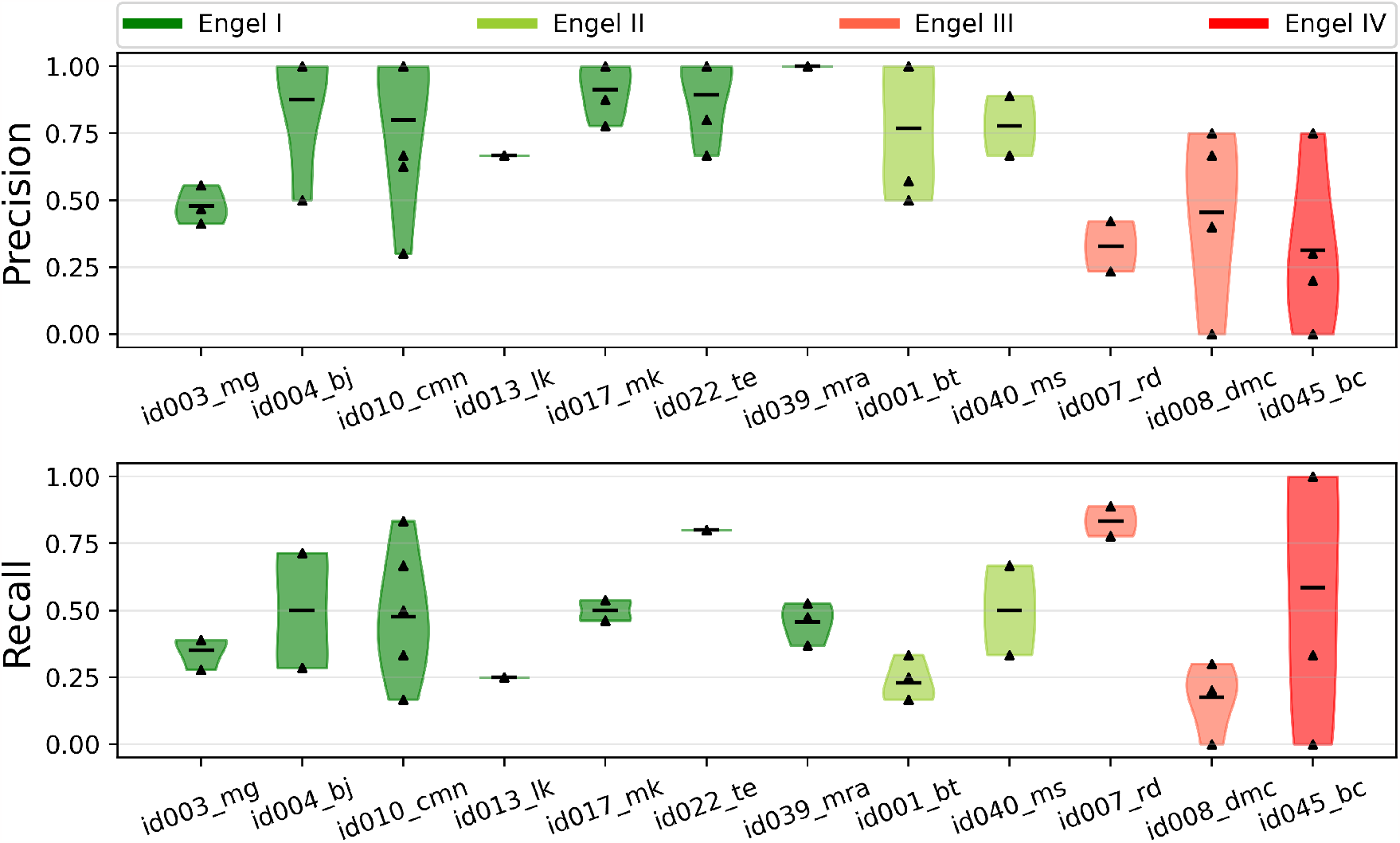
Analysis of the effect of mapping resection mask from VEP parcellation level to high resolution on inference accuracy, specifically on recall. Similar to Fig.4a, except precision/recall are computed at parcellation level. Resection masks at high resolution are constructed from the post surgical resection masks available at parcellation level. It’s effect on recall is studied by mapping the predicted EZ to parcellation level and computing precision and recall by comparing against resection masks at the parcellation level itself.

**Figure 6:**
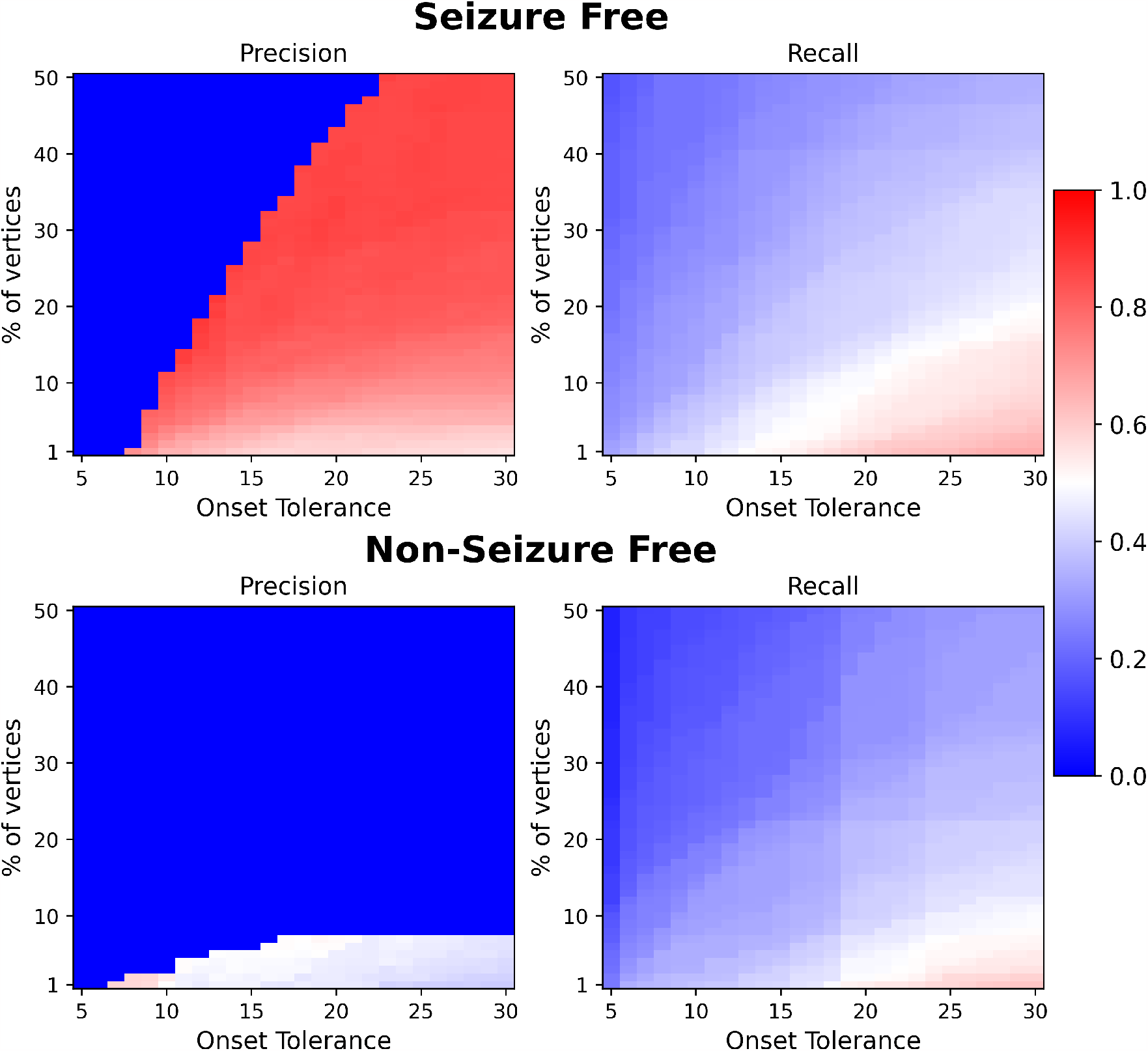
Analysis of effect of various thresholds performed in mapping EZ to parcellation level on prediction accuracy. Predicted EZ at high resolution is mapped to parcellation level by thresholding on the minimum percentage of vertices in a region and the seizure onset time to be considered as part of EZ. Average precision and recall, averaged across patients, across different values of these thresholds in seizure free group (top) and non-seizure free group (bottom). Note that in computing precision there can be no regions in the EZ for low values of onset tolerance and high values of percentage of vertices. This can lead to division by zero errors, hence, if such an error occurred for any of the patients at a particular threshold then the precision is set to zero.

Identification of seizure onset zone using purely data driven signal analysis of SEEG time series such as EI, EM and ER [2, 3, 5] are by construction spatially constrained to identify EZ within the implanted regions. Similar to previous VEP studies [16, 18, 17, 31], the generative model used in this study also includes a forward model, the so called the gain matrix, transforming latent source activity to the SEEG time series. The forward model could account for seizure activity originating in regions beyond the implanted regions on the observed SEEG. The efficacy of the forward model in discovering EZ beyond implanted regions is studied by analyzing the number of regions in the predicted EZ that are beyond the implanted regions. Regions within a radius of 3 mm of any sensor are considered to be implanted. The number of regions, averaged across patients in each group, in the predicted EZ not part of the implantation is shown in Fig.7a. In seizure free group 25% of the regions are found to be outside implantation while in the non-seizure free group it is 38%. Previous results (Fig.4, 5) demonstrated that there are more false positives in non-seizure free group compared to the seizure free group. Fig.7a also shows the percentage of predicted regions in EZ that are outside implantation. Interestingly, among the false positives the non-seizure free group showed much higher regions outside implantation compared to seizure free group. Whereas, among the true positives the difference is much smaller. It is reasonable to assume that among the non-seizure free patients the implantation missed some the EZ. These findings provide confidence to that effect and demonstrates the efficacy of the proposed method in identifying EZ beyond the implanted regions. However, under such rational, it is also reasonable to hypothesize that: (i) among seizure free patients implantation should have covered the EZ and (ii) the true positives should be covered by the implantation irrespective of the group. Fig.7b shows the percentage of predicted EZ regions not implanted as the radius from sensor is increased from 1 mm to 10 mm. As hypothesized, as the radius is increased, all the regions in predicted EZ of seizure free patients are covered by the implantation and among the true positives the percentage of regions outside implantation have reduced to less than 2% in both groups. The distributions of percentage of predicted regions in EZ which are not implanted across all patients and seizures are shown in Fig.7c.

**Figure 7:**
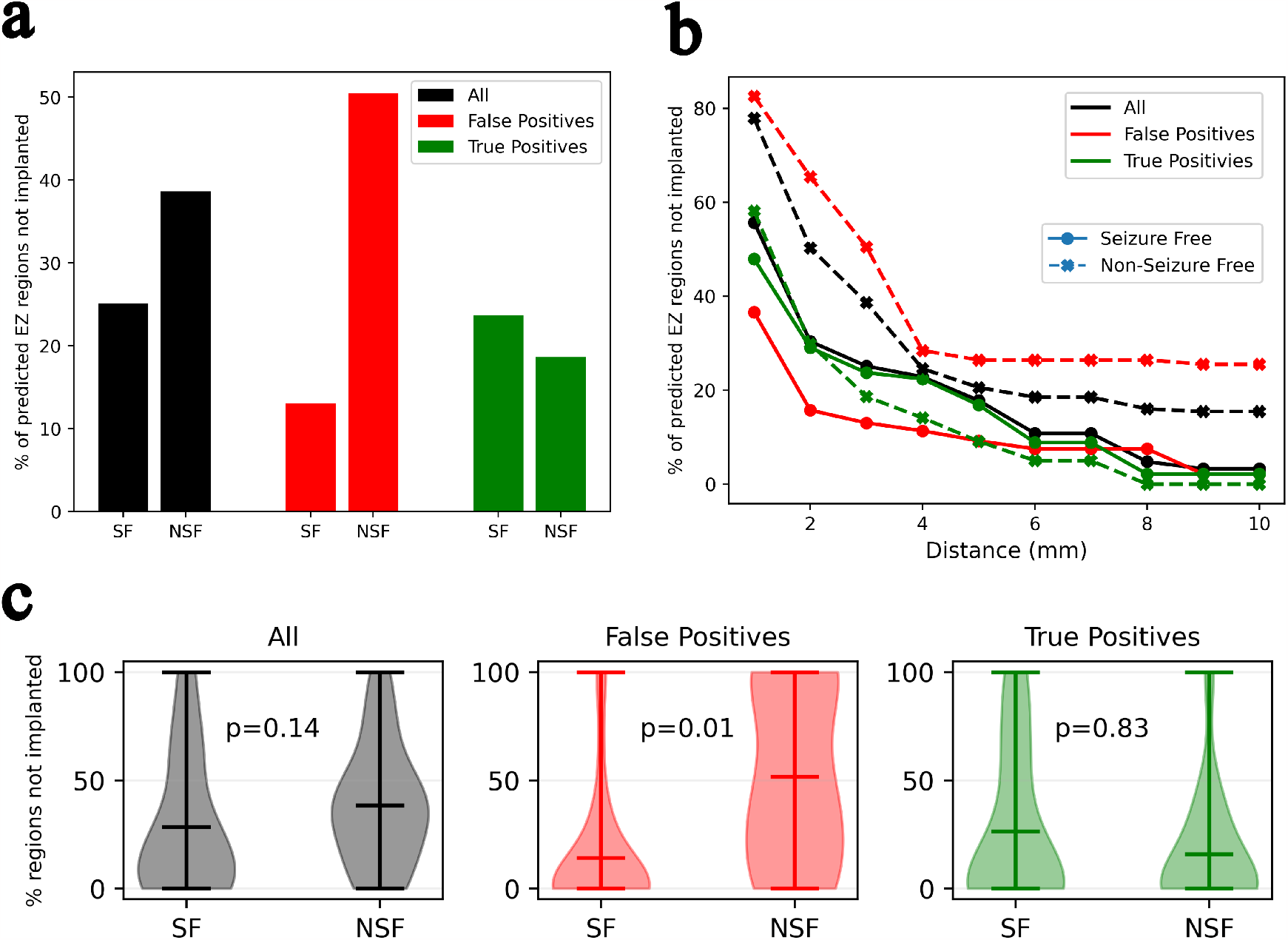
EZ discovery beyond implanted regions. (a) Comparison of percentage of regions, averaged across patients, beyond the implanted regions in seizure free and non-seizure free groups. The tolerance for considering a region as implanted is set to 3 mm, i.e. all regions within a radius of 3 mm of any implanted sensor are considered to be part of implanted regions. (b) Percentage of regions beyond implantation as the distance tolerance defining implantation is increased from 1 mm to 5 mm. (c) Distribution of percentage of regions not implanted across all patients and seizures with implantation radius of 3 mm.

### Comparison to Neural Mass Models

Lastly, the accuracy of the neural field extension of the VEP (VEP-NFM) framework proposed in this study is compared against the previous VEP approaches with neural mass model (VEPNMM) [18, 31] against the same retrospective dataset. In the seizure free group, at an onset tolerance of 10 seconds, the number of false positives have drastically reduced as can be seen from the precision in Fig.8a,b. A paired samples t-test was performed to compare precision and recall between VEP-NMM and VEP-NFM. There is a statistically significant difference in precision between VEP-NFM (M=0.85, SD=0.21) and VEP-NMM (M=0.56, SD=0.34); t(28)=5.41, p¡0.01. For recall, there is not a significant difference between VEP-NFM (M=0.25, SD=0.21) and VEP-NMM (M=0.29, SD=0.27); t(28)=-1.48, p=0.15. In the non seizure free group, there is not a statistically significant difference between VEP-NMM and VEP-NFM in either precision or recall. At an onset tolerance of 30 seconds, precision is found to be statistically significant in both seizure free and non-seizure groups, while, recall is not. Average precision and recall in VEP-NMM and VEP-NFM across as the onset tolerance threshold is increased from 5 to 30 seconds is shown in Fig.8.

**Figure 8:**
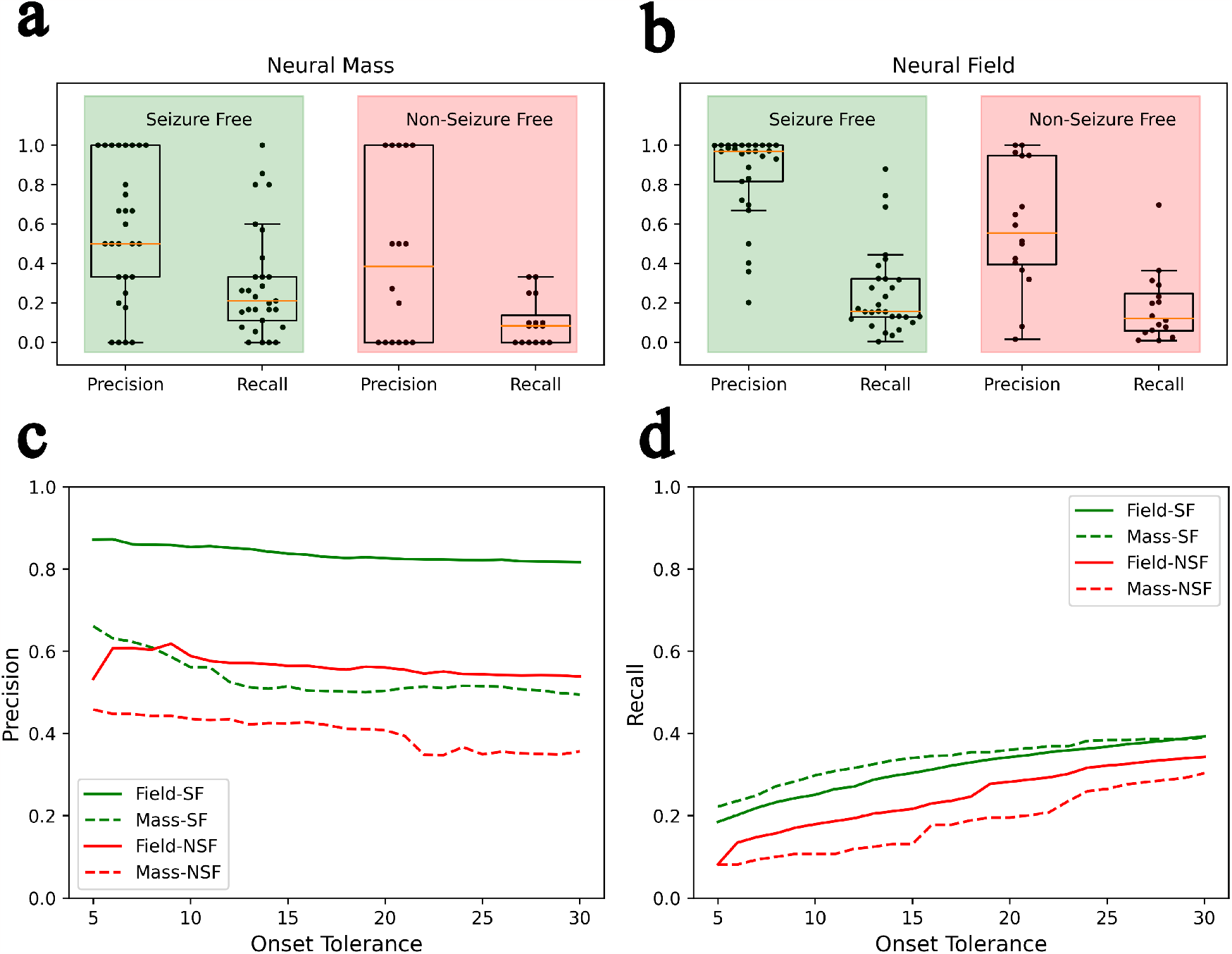
Comparison of prediction accuracy of VEP with neural mass and neural fields model. (a) Precision and recall of VEP with neural mass 2D Epileptor model (VEP-NMM) against a retrospective dataset (b) Precision and recall of VEP with neural field 2D Epileptor model (VEP-NFM) against the same retrospective dataset. (c,d) Precision and recall of VEP-NMM and VEP-NFM across various onset tolerance thresholds.

## Discussion

In this study, we explored the development of personalized models for the identification of EZ by extending the VEP approach [18, 17, 31] to a neural field model. Our primary objective was to enhance the spatial resolution of the predicted EZ using a neural field model, which theoretically offer infinite spatial resolution. We employed a recently proposed pseudo spectral method [33] to approximate solutions to the neural field equations, thereby avoiding the computational challenges associated with simulation and model inversion of neural field models.

Our findings demonstrate the success of our approach in achieving improved spatial resolution for EZ prediction. These findings could play a important role in the context of epilepsy research, as the accurate localization of the EZ is critical for guiding surgical interventions and improving patient outcomes. The use of neural field models proved to be a fruitful advancement, allowing us to harness their potential for high spatial precision. The adoption of the pseudo spectral method reduced the computational cost of model inversion, providing a practical solution to overcome the computational hurdles that often accompany neural field models. This method presents a promising avenue for future research and may find applications in other computational neuroscience domains requiring model inversion.

Systematic testing against synthetic data revealed that the prediction accuracy of our model remained robust against variations in spatial resolution, demonstrating its reliability in settings where data quality and resolution may vary. However, we noted a degradation in accuracy with higher degrees of spherical harmonics. This observation prompts further investigation into the trade-offs between spatial resolution and computational efficiency in our approach. Strategies to mitigate this effect could be explored in future work.

The validation against retrospective data yielded encouraging results. Our model predictions exhibited a strong match with seizure-free patients, underscoring its potential as a tool for aiding in the diagnosis and treatment planning for epilepsy patients. Conversely, a notable mismatch was observed with non-seizure-free patients, highlighting the model’s potential for identifying individuals at a higher risk of recurrent seizures. These clinical insights demonstrate the practical applicability of our research and the potential for improved patient outcomes.

We acknowledge several limitations in our study. The retrospective dataset used for validation is small and thereby validation against larger datasets is needed. Future research should consider more comprehensive datasets to enhance confidence of the proposed approach. Model inversion is performed using MAP which provides point estimates using an optimization algorithm. Hence the drawbacks of an optimization algorithms such as getting stuck in a local minima also apply to our results. One common technique to partially mitigate this challenge is to run multiple optimizations with different initial conditions, which we did employ in this study. To address issues with MAP, we are currently exploring variational inference using more sophisticated density approximation methods such as Masked auto-regressive flows [41] and Free-form Jacobian of reversible dynamics (FFJORD) [42].

Moving forward, our research paves the way for several promising directions. Clinical validation through prospective studies is essential to further assess the reliability and real-world applicability of our models. We also anticipate opportunities for algorithm refinement, focusing on strategies to address the degradation in accuracy associated with higher degrees of spherical harmonics. Finally, the integration of our approach with other computational neuroscience techniques and complementary data sources could enhance the accuracy and clinical relevance of our personalized models.

In conclusion, our study advances the VEP framework by offering a novel approach to personalized models of epileptic seizure propagation. The use of neural field models and the pseudo spectral method has shown promise in improving spatial resolution for EZ prediction, with clinical implications for diagnosis and treatment planning. While challenges and limitations remain, our research opens exciting avenues for future investigations.

## Acknowledgments

This research was supported by the Fondation pour la Recherche Médicale (Grant DIC20161236442), European Union’s Horizon 2020 research and innovation programme under grant agreement No. 945539 (SGA3) Human Brain Project, and the SATT Sud-Est (TVB-Epilepsy). This work has been carried out within the Fédération Hospitalo-Universitaire EPINEXT with the support of the Recherche Hospitalo-Universitaire EPINOV (Grant ANR-17-RHUS-0004) funded by the “Investissements d’Avenir” French Government program managed by the French National Research Agency (ANR).

## Data availability

The patient data sets cannot be made publicly available due to the data protection concerns.

## Author Contributions

ANV, VJ and MMW designed the study. ANV implemented the code, performed analysis and prepared the manuscript. FB provided the retrospective data. JD preprocessed the retrospective data using the reconsturction pipeline and provided the high resolution surfaces, gain matrices and connectomes. DD helped understanding the details of pseudospectral methods. All authors reviewed and edited the manuscript.

