## Supplementary Material for "Improving epileptogenic zone estimation using Bayesian inference on neural field models"

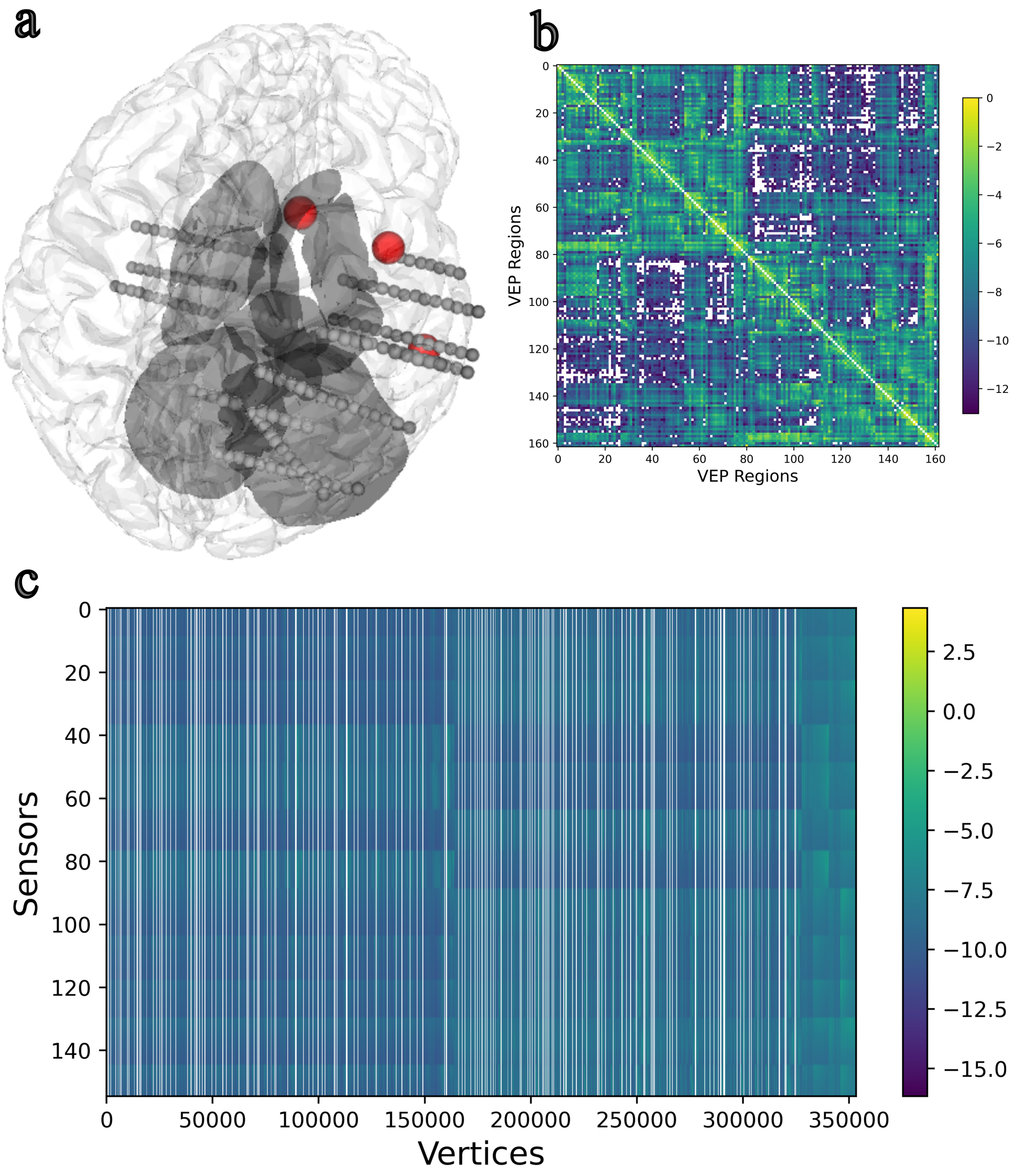


Figure 1: Details of patient $id004$ used for generating synthetic data. (a) SEEG electrode implantation. Sensors are shown in gray solid balls and centers of the brain regions in Epileptogenic Zone are shown in red balls. (b) Structural Connectome constructed from diffusion MRI. (c) Gain matrix

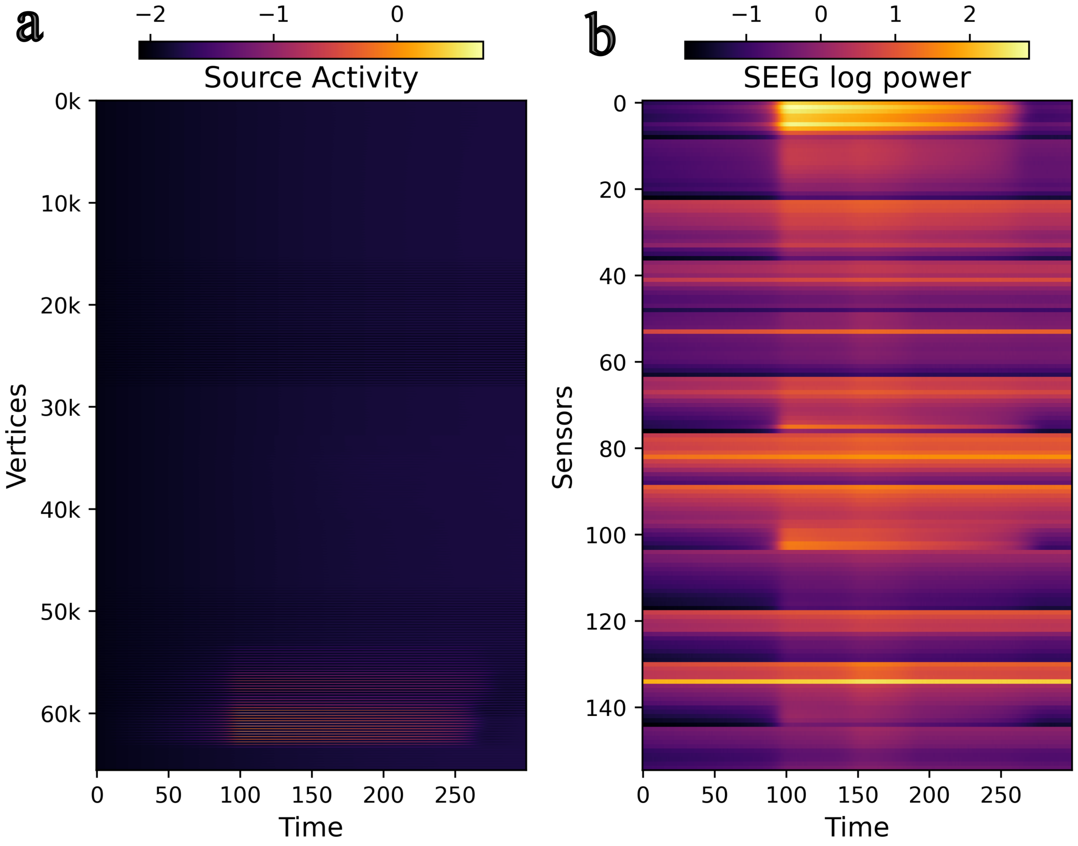


Figure 2: Synthetic data generated based on data of patient *id*004 (a) Simulated source activity (b) Observed SEEG log power obtained by projecting the simulated source activity in "a" using the gain matrix shown in 1c

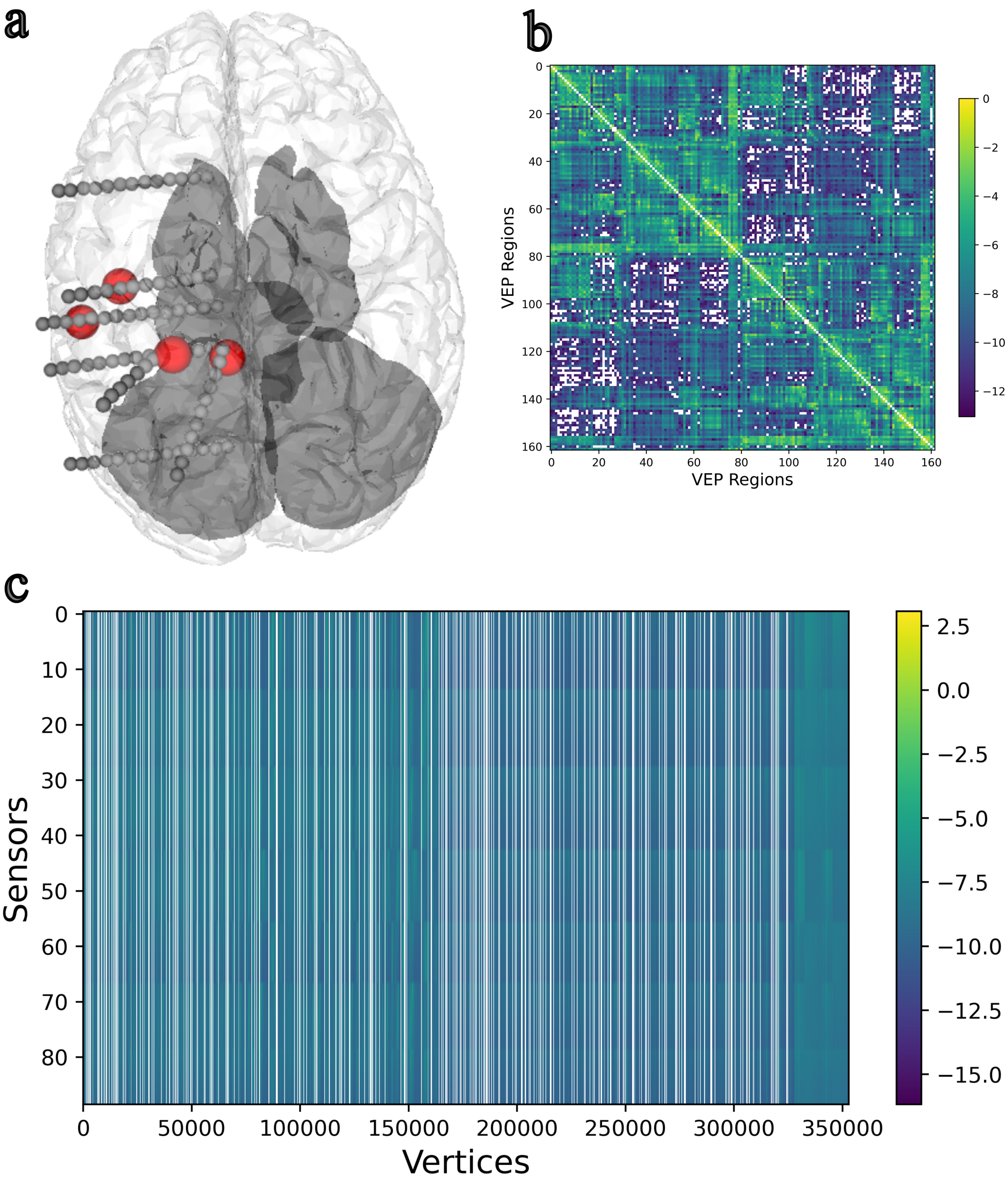


Figure 3: Details of patient $id022$ used for generating synthetic data. (a) SEEG electrode implantation. Sensors are shown in gray solid balls and centers of the brain regions in Epileptogenic Zone are shown in red balls. (b) Structural Connectome constructed from diffusion MRI. (c) Gain matrix

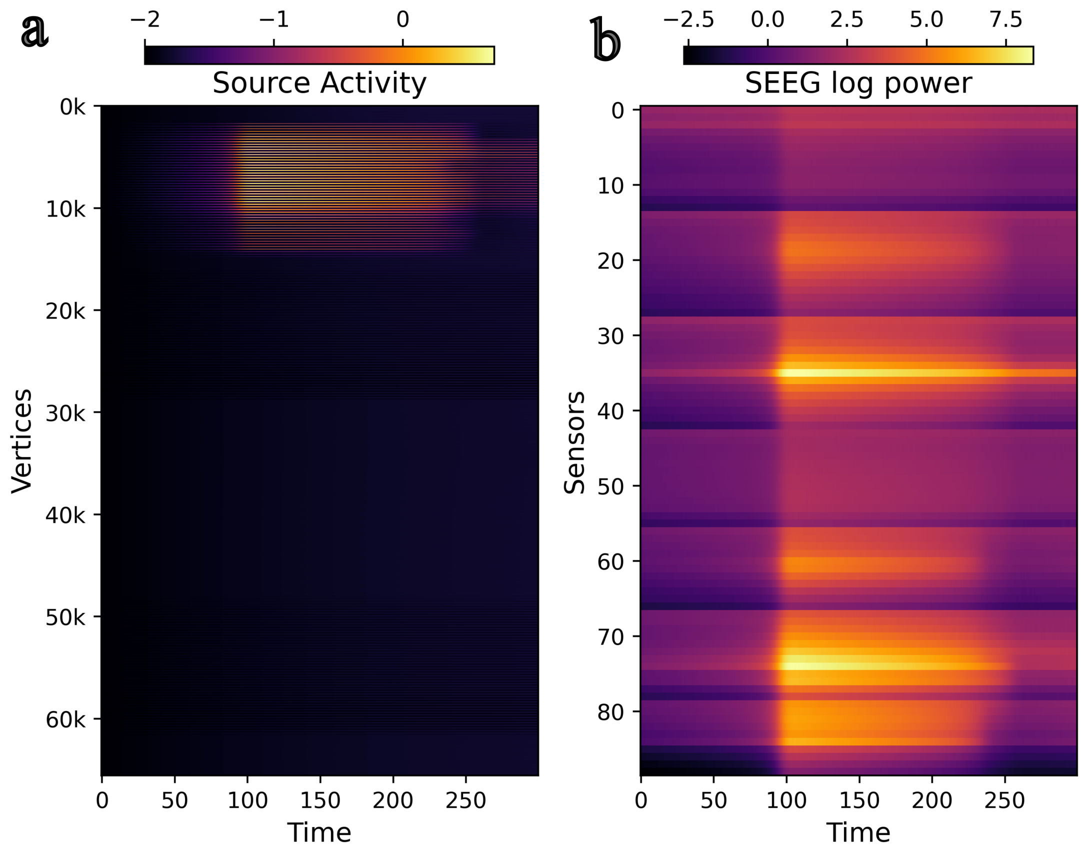


Figure 4: Synthetic data generated based on data of patient *id*004 (a) Simulated source activity (b) Observed SEEG log power obtained by projecting the simulated source activity in "a" using the gain matrix shown in 3c

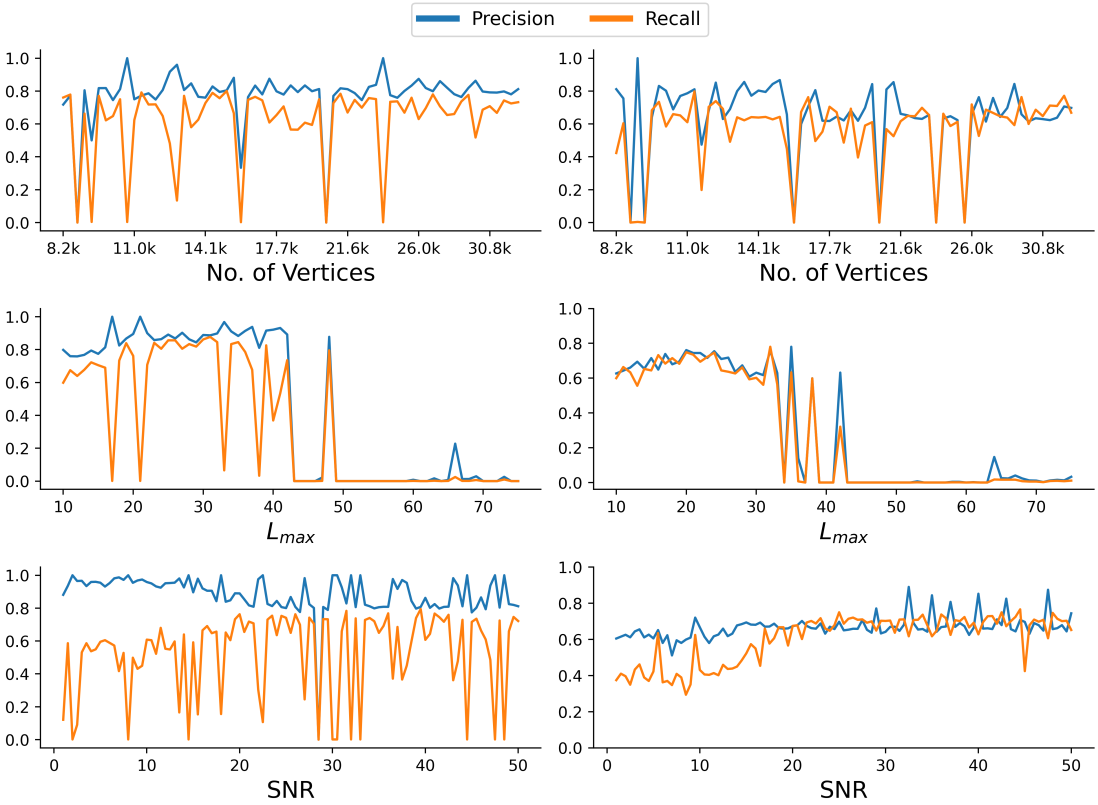


Figure 5: Accuracy of inferred EZ in patient id004 across different parameter sweeps. (a) Accuracy across different spatial resolutions (b) Accuracy across different mode truncation (c) Accuracy across different signal to noise ratio.
